# Practical strategies for SARS-CoV-2 RT-PCR testing in resource-constrained settings

**DOI:** 10.1101/2021.02.18.21251999

**Authors:** Meredith S. Muller, Srijana Bhattarai Chhetri, Christopher Basham, Tyler Rapp, Feng-Chang Lin, Kelly Lin, Daniel Westreich, Carla Cerami, Jonathan J. Juliano, Jessica T. Lin

**Affiliations:** Institute of Global Health and Infectious Diseases, University of North Carolina School of Medicine, Chapel Hill, NC USA; MRC Unit The Gambia at the London School of Hygiene & Tropical Medicine; Gillings School of Global Public Health, University of North Carolina, Chapel Hill, NC USA

## Abstract

**Backgroun:** Standard nasopharyngeal swab testing for SARS-CoV-2 detection by PCR is not always feasible due to limitations in trained personnel, personal protective equipment, swabs, PCR reagents, and access to cold chain and biosafety hoods.

**Method:** We piloted the collection of nasal mid-turbinate swabs amenable to self-testing, including both standard polyester flocked swabs as well as 3D printed plastic lattice swabs, placed into either viral transport media or an RNA stabilization agent. Quantitative SARS-CoV-2 viral detection by RT-qPCR was compared to that obtained by nasopharyngeal sampling as the reference standard. Pooling specimens in the lab versus pooling swabs at the point of collection was also evaluated.

**Results:** Among 275 participants, flocked nasal swabs identified 104/121 individuals who were PCR-positive for SARS-CoV-2 by nasopharyngeal sampling (sensitivity 87%, 95% CI 79-92%), mostly missing those with low viral load (<10^3 viral copies/uL). 3D-printed nasal swabs showed similar sensitivity. When nasal swabs were placed directly into an RNA stabilizer, the mean 1.4 log decrease in viral copies/uL compared to nasopharyngeal samples was reduced to <1 log, even when samples were left at room temperature for up to 7 days. Pooling sample specimens or swabs both successfully detected samples >10^2^ viral copies/uL.

**Conclusions:** Nasal swabs are likely adequate for clinical diagnosis of acute infections to help expand testing capacity in resource-constrained settings. When collected into an RNA preservative that also inactivates infectious virus, nasal swabs yielded quantitative viral loads approximating those obtained by nasopharyngeal sampling.

## BACKGROUND

Since the start of the SARS-CoV-2 pandemic, testing has been a cornerstone of the public health response. The de facto standard for clinical testing is PCR from nasopharyngeal (NP) swabs. However, nasopharyngeal sampling must be performed by trained staff using personal protective equipment (PPE). Shortages in both, as well as NP swabs themselves, often manifest when case counts climb. A wide array of strategies amenable to self-collection have been piloted to expand testing capacity, including the collection of nasal swabs, oropharyngeal and tongue swabs, saliva, and oral rinses (1–3). The volume of tests conducted can also become burdensome and lengthen turnaround time, spurring interest in pooled testing strategies in low prevalence and settings (4–8). Finally, regarding sample storage and transport, viral specimens are typically placed in viral transport medium, and CDC recommends maintenance of cold chain prior to processing (1), but this may not be possible in all settings.

In order to implement a household transmission study in the early phases of the epidemic in North Carolina, when shortages of PPE and swabs were prevalent, we adopted a strategy of self-collected nasal swabs from household members during follow-up. Here we compare this strategy to concurrently collected nasopharyngeal swabs at enrollment in our study population. We piloted different types of swabs stored in different media. Given interest in pooling strategies for high throughput testing, we also used our cohort to test two different pooling strategies: pooling swabs at the point of collection or pooling sample lysate in the lab. Our findings provide confidence in using self-collected nasal swabs, preferably stored in an RNA stabilizer, when nasopharyngeal sampling is not feasible.

## METHODS

### Clinical samples

Clinical samples were collected as part of a SARS-CoV-2 household transmission study conducted in the Piedmont region of North Carolina. The study received ethical approval from the Institutional Review Board at the University of North Carolina-Chapel Hill and is registered at clinicaltrials.gov (NCT04445233). Participants were enrolled if they were adults that tested positive for SARS-CoV-2 by PCR at the UNC Respiratory Diagnostic Center and shared a living space with one or more persons who also agreed to participate. At enrollment, a standard clinician-collected nasopharyngeal (NP) swab was performed, followed by up to two other nasal swabs (on different sides) that were either collected by study staff or self-collected by the participant or their guardian with guidance from study staff (**Figure S1**). For nasal sampling, participants were instructed to insert the swab about 1-2 inches into one nostril, then swirl 5 times while slowly withdrawing the swab before placing it into the collection tube. All samples were placed into a cooler on ice prior to processing in a BSL2+ laboratory space.

### Sample collection strategies

Flocked NP swabs were collected into 3mL of Becton Dickinson’s co-packaged universal viral transport system. Two types of nasal swabs designed for mid-turbinate sampling (NMT) were used: flocked NMT swabs (COPAN, Murrietta CA) and 3D-printed lattice NMT swabs (Resolution Medical, Fridley MN) (**Figures S1**). Both were collected into 3mL viral transport media (VTM) prepared using CDC SOP# DSR-052-05. Upon sample receipt in the laboratory, 1mL of the collected sample was combined with 1mL 2X DNA/RNA Shield, a nucleic acid preservation agent and lysis buffer (Zymo Research), and stored at -80°C until extraction. RNA was extracted from 200uL of the lysate using the Quick-RNA Viral 96 Kit (Zymo Research) and eluted in 20uL of water. We also evaluated the effect of storage media by collecting flocked NMT swabs directly into 3mL of 1X DNA/RNA Shield (Shield), with aliquots either frozen immediately upon return to the lab or left at room temperature for 4 or 7 days before being stored at -80°C. RNA was extracted from 100 uL of the lysate using the same extraction and elution protocols.

### qRT-PCR viral quantification

Samples were tested using a CDC RT-qPCR protocol authorized for emergency use that consists of three unique assays: two targeting regions of the virus’ nucleocapsid gene (N1, N2) and one targeting human RNase P gene (RP) (Catalog # 2019-nCoVEUA-01, Integrated DNA Technologies) (9). 5uL of extracted RNA was added to 15uL of each assay’s reaction mixture containing TaqPath 1-Step RT-qPCR Master Mix, CG (Thermofisher Scientific) and the corresponding primer-probe set (IDT), followed by the recommended thermocycler protocol. Plasmid DNA containing the human RPP30 gene and SARS-CoV-2 in vitro transcribed RNA control (nCoVPC, IDT) were used as positive controls. Water was used as a negative extraction control. Samples were designated positive if all three PCRs were positive (N1 and N2 for virus, RP for adequate sampling). If the N1 and N2 PCRs were negative, but the RP assay had a Ct value ≥30 or was negative, suggesting inadequate sampling, then the sample was re-extracted. The second result was reported if the RP Ct value was <30 or if both N1 and N2 PCRs were positive regardless of RP Ct value.

The viral load of each sample, in copies/uL, was extrapolated from standard curves generated for each viral assay (N1 and N2) using serial dilutions of nCoVPC (2 to 100,000 viral RNA copies/uL). The average copies/uL between the N1 and N2 assays was used as the final quantitative viral load. Based on the sample collection and RNA extraction volumes as well as volume of template RNA used in the RT-qPCR (5uL), this viral load represents the number of viral RNA copies per 5 uL of VTM or Shield sample.

### Pooling strategies

The efficacy of pooling NMT samples was examined through two different approaches: pooling swabs at the point of care into the same collection vessel and pooling individual sample lysates prior to extraction. For the first strategy, self-collected 3D-printed lattice NMT swabs from each member of a household of three or more were collected and pooled together in 5mL of VTM. This was done at one or more of the study visits for each household. 200uL of the sample lysate was extracted and quantified as above. Results were compared to the self-collected individual flocked NMT swab collected at the same visit. In the second pooling strategy, one qRT-PCR positive sample lysate from a flocked NMT swab (pre-RNA extraction) was pooled with sample lysate from negative individuals to construct pool sizes of 5, 10, 15, and 20. The Ct values of twelve samples with viral copies/uL ranging from 10^1^ to 10^7^ were compared to the Ct values of their corresponding pools.

### Statistical analysis

A probit analysis of results from the nCoVPC plasmid control concentrations (ranging from 2 to 100,000 copies/uL as part of standard curves generated in every RT-qPCR run) by parametric curve fitting to hit rate data was used to determine the limit of detection (LOD) of the N1 and N2 qRT-PCR assays. Samples that were positive in both N1 and N2 assays, but with an average viral load that fell below the LOD were categorized as indeterminate. The sensitivity and specificity of different swab types for RT-qPCR detection of SARS-CoV-2 was calculated using flocked NP swabs as the reference standard. Additionally, the difference in the quantitative viral load was compared for different collection strategies. Comparisons were made on the log scale and analyzed using Wilcoxon matched-pairs signed rank testing with a p-value<0.05 considered significant. Statistical analyses were performed using GraphPad Prism 8 and SAS 9.4 (Cary, NC).

## RESULTS

We report data from 644 swab samples collected from 275 participants (91 households) at enrollment, 24 pools collected at follow-up or enrollment, and 44 pools constructed from participant samples in the lab. Participants ranged in age from 1-77 years old, with 71% >18 years of age.

### Limit of detection of RT-qPCR assay

Probit analysis of nCoVPC plasmid control concentrations tested in 33 RT-qPCR runs yielded a limit of detection (LOD) for the N1 and N2 assays of 9 and 13 copies/uL, respectively (**Table S1**). The average LOD between the two assays, 11 copies/uL, was used as the cutoff for sample positivity. A sample was deemed positive if the average viral load derived from the cycle threshold (Ct) values of N1 and N2 corresponded to a concentration ≥11 copies/uL, indeterminate if <11 copies/uL, and negative if either assay failed to amplify. Altogether, 21/702 (3.0%) samples tested fell into the indeterminate category. Another 33 (4.7%) samples only amplified in one assay (N1 or N2 assay), but with a Ct value corresponding to a viral load that fell below the LOD. Only 2 samples (0.3%) were discordant between the N1 and N2 assays (positive in one but not the other).

### Comparison of collection swabs and storage medium

Compared to nasopharyngeal sampling, flocked nasal mid-turbinate (NMT) swabs displayed slightly decreased sensitivity, but were well-accepted by the participants and yielded adequate sampling. Altogether, at enrollment, 275 study participants completed 226 NP swabs and 418 NMT swabs (255 flocked and 51 3D-printed in VTM, 112 flocked in Shield) (**Figure S1**). Of the 49 participants that declined to do NP swabs, 46 agreed to at least one type of NMT swab. Inadequate sampling, as defined by negative N1 and N2 PCRs in concert with a negative human RP PCR or Ct ≥30, occurred in small numbers of flocked swabs, but a substantial proportion of 3D-printed plastic lattice swabs: 1/226 (0.4%) of NP swabs, 14/343 (4.1%) of flocked NMT swabs, and 11/51 (21.6%) of 3D-printed plastic lattice swabs.

Using NP swabs as the reference standard, flocked NMT swabs showed excellent specificity (98%, 95% CI 90-100%) but slightly decreased sensitivity (87%, 95% CI 79-92%) for SARS-CoV-2 detection by RT-qPCR (**Figures 1&2**). Of 173 NP-NMT swab pairs, 104 were both positive, 52 both negative, and 10% (17/173) were discordant. Three of these discordances were likely due to inadequate sampling (1 NP, 2 NMT swabs with RP Ct value ≥30), while 71% of the rest (10/14) occurred in samples with low viral loads (<10^3^ viral copies detected in the NP swab). In the 104 positive swab pairs, NMT samples displayed lower average viral loads (Spearman correlation coefficient=0.67, **Figure 1**), with a mean 1.3 log decrease in viral copies/uL (IQR 0.6 - 2.1 log viral copies/ul) compared to NP sampling (p<0.0001) (**Figure 3A**). This was at least partly due to a sampling difference, as NMT swabs also showed on average 3.1 cycles higher Ct values in the human RP PCR (**Figure S2**).

**Figure 1.**
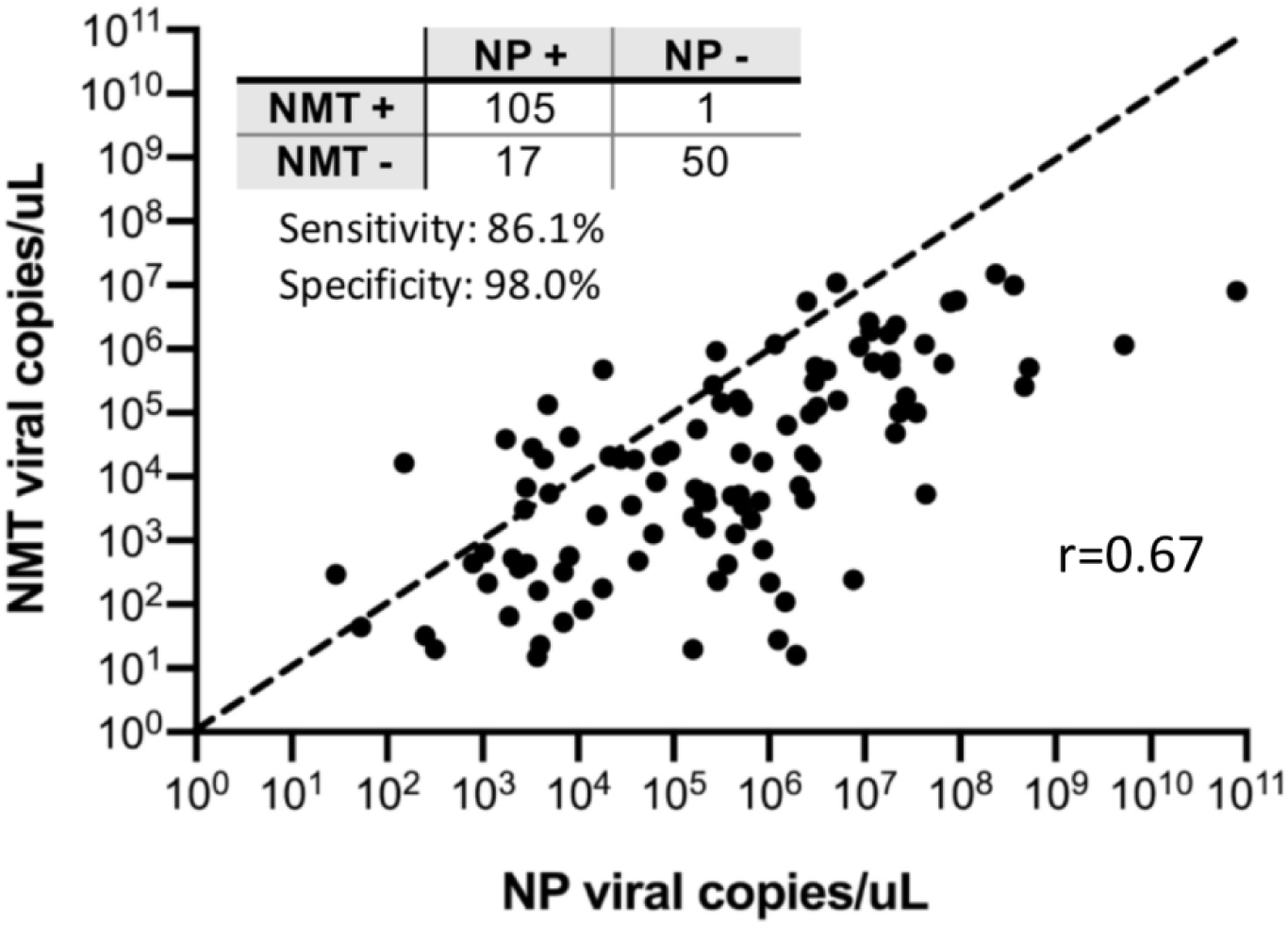
Concordance and comparison of SARS-CoV-2 viral loads from paired nasopharyngeal (NP) and nasal mid-turbinate (NMT) swabs. Paired NP and NMT swabs from 173 participants showed overall good concordance, with most discordances (15/16) arising from positive NP/negative NMT samples. Quantitative viral loads derived from the average of N1 and N2 qRT-PCR assays favored NP swabs compared to NMT swabs. A y=x dashed line is drawn for reference.

**Figure 2.**
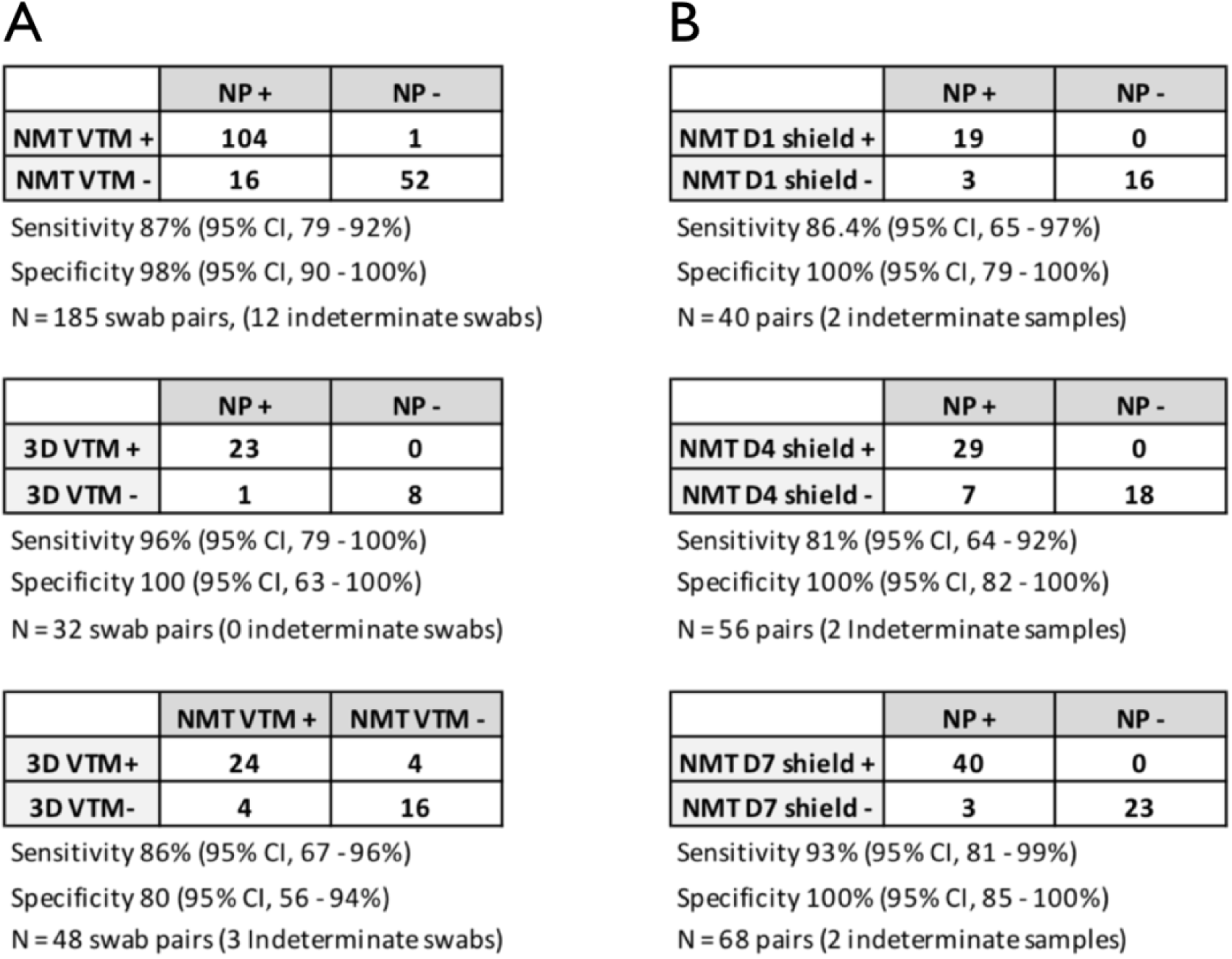
Concordance of SARS-CoV-2 RT-qPCR detection between nasopharyngeal (NP) swabs and two different nasal mid-turbinate (NMT) swab types, stored in viral transport media (VTM) or 1x DNA/RNA shield (Shield). In (A), sensitivity and specificity of standard flocked (NMT) or 3D-printed (3D) nasal swabs collected into VTM are shown using NP swabs with co-packaged universal viral transport system as the reference standard. Concordance of flocked vs. 3D nasal swabs is also shown. In (B), flocked NMT swabs were stored in Shield, and sample aliquots were directly frozen on day 1 (D1) or kept at room temperature before being stored at -80C on day 4 (D4) or day 7 (D7). Note that samples with indeterminate viral load (<11 copies/ul) were not included in the sensitivity/specificity analyses.

**Figure 3.**
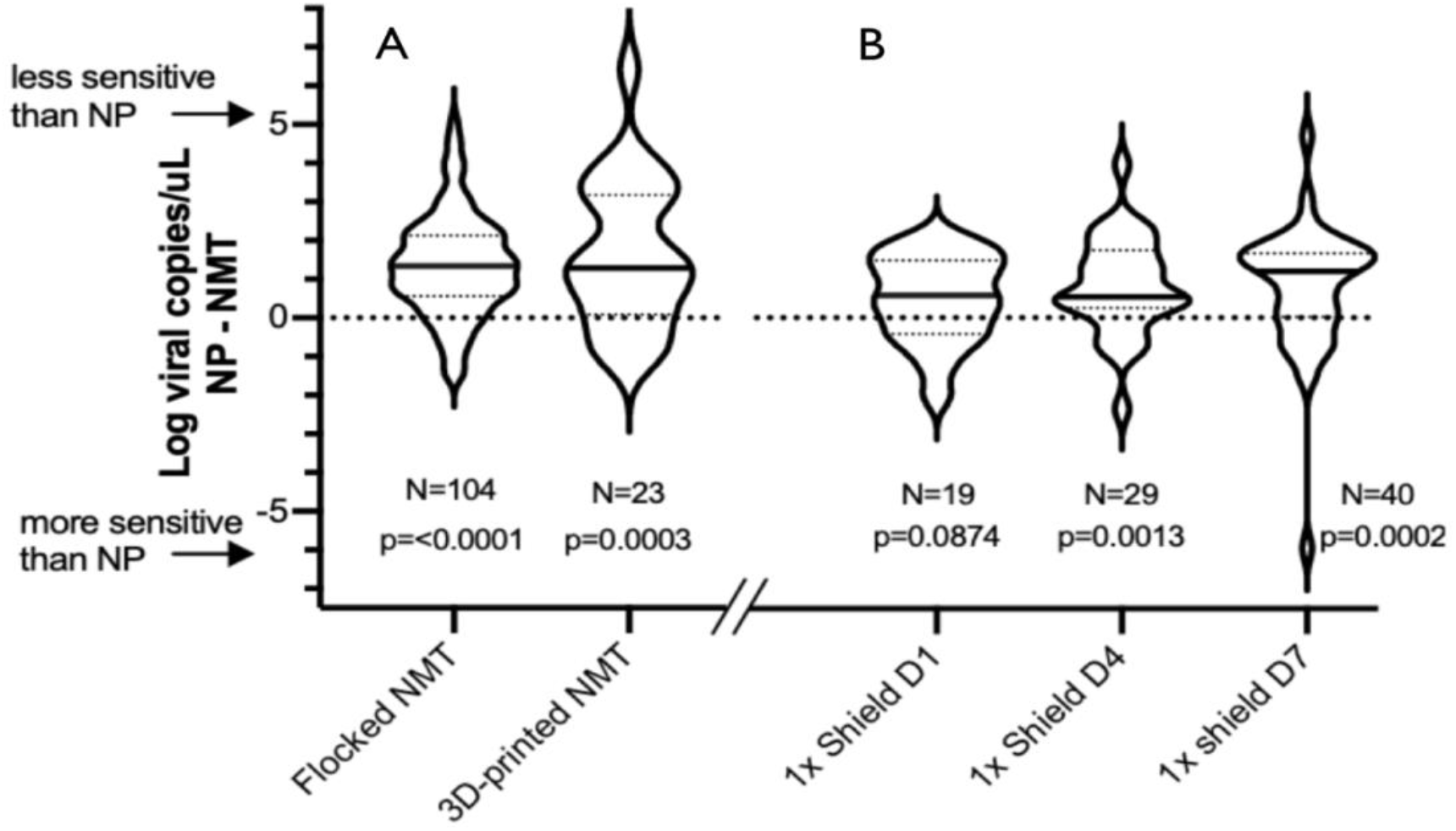
Comparison of SARS-CoV-2 viral loads between standard NP swabs and NMT or 3D NMT swabs (A) as well as NMT swabs collected into 1x DNA/RNA Shield and stored for different intervals (B). The distribution of the difference in log viral load is depicted for each comparison. Median log-fold changes are indicated by a solid line with interquartile values indicated by dotted lines. The number of sample pairs is indicated for each comparison.

Though the 3D-printed plastic lattice NMT swabs were more likely to lead to inadequate sampling, positive samples showed quantitative viral loads similar to flocked NMT swabs (**Figure 3A**). This was true despite on average 1.2 higher Ct values for the human RP assay in the 3D vs. flocked swabs. Compared to NP sampling, 3D-printed NMT swabs displayed 95.7% sensitivity (95% CI 78.1%-99.9%) and 100% specificity (95% CI 63.1-100%) among 48 swab pairs (**Figure 2**).

Placing flocked NMT swabs directly into 1x DNA/RNA Shield did not improve the sensitivity of detection, but did result in viral loads comparable to those obtained by NP sampling. Aliquots of Shield samples were either directly stored at -80C (similar to other samples collected on day 1), or left out at room temperature for 4 or 7 days prior to freezing and processing. All NMT Shield samples showed a specificity of 100% compared to NP swabs, while sensitivity ranged 86%, 78%, and 91% for the samples frozen at day 1, 4, and 7, respectively (**Figure 2**).

Altogether, regardless of how many days the Shield samples were left out, the overall sensitivity was 85% (95% CI 77-92%). While sensitivity for detection was slightly diminished, quantitative viral loads derived from NMT Shield aliquots frozen on day 1 were comparable to NP viral loads (mean decrease of 0.5 log viral copies/uL (IQR -0.3-1.4), p=0.09) (**Figure 3B**). For aliquots left at room temperature until day 4 and day 7, we observed a mean decrease that was still <1 log viral copies/uL compared to NP sampling (mean 0.8 and 0.8 log viral copies/ul, respectively (p=0.001 and p=0.0002) (**Figure 3B)**.

### Pooling strategies

The pooling strategies implemented were sufficient for detecting samples with viral loads >10^2 copies/uL but were not as sensitive as individual swabs for detecting samples with lower viral loads. Of the 24 pools of 3D-NMT swabs pooled at the point of care, 3 were indeterminate, and 2 (8%) yielded discordant results (depicted as red stars in **Figure 4**). Under the assumption that the concurrently collected individual flocked NMT swabs were accurate, the two discordant results were false negative pools where the individual swab had a viral load <100 copies/uL, close to the limit of detection (**Table S2**). Of the 22 concordant pools, 8 were negative and 11 were positive, mostly with individual swab viral loads ≥10^2 copies/uL.

**Figure 4.**
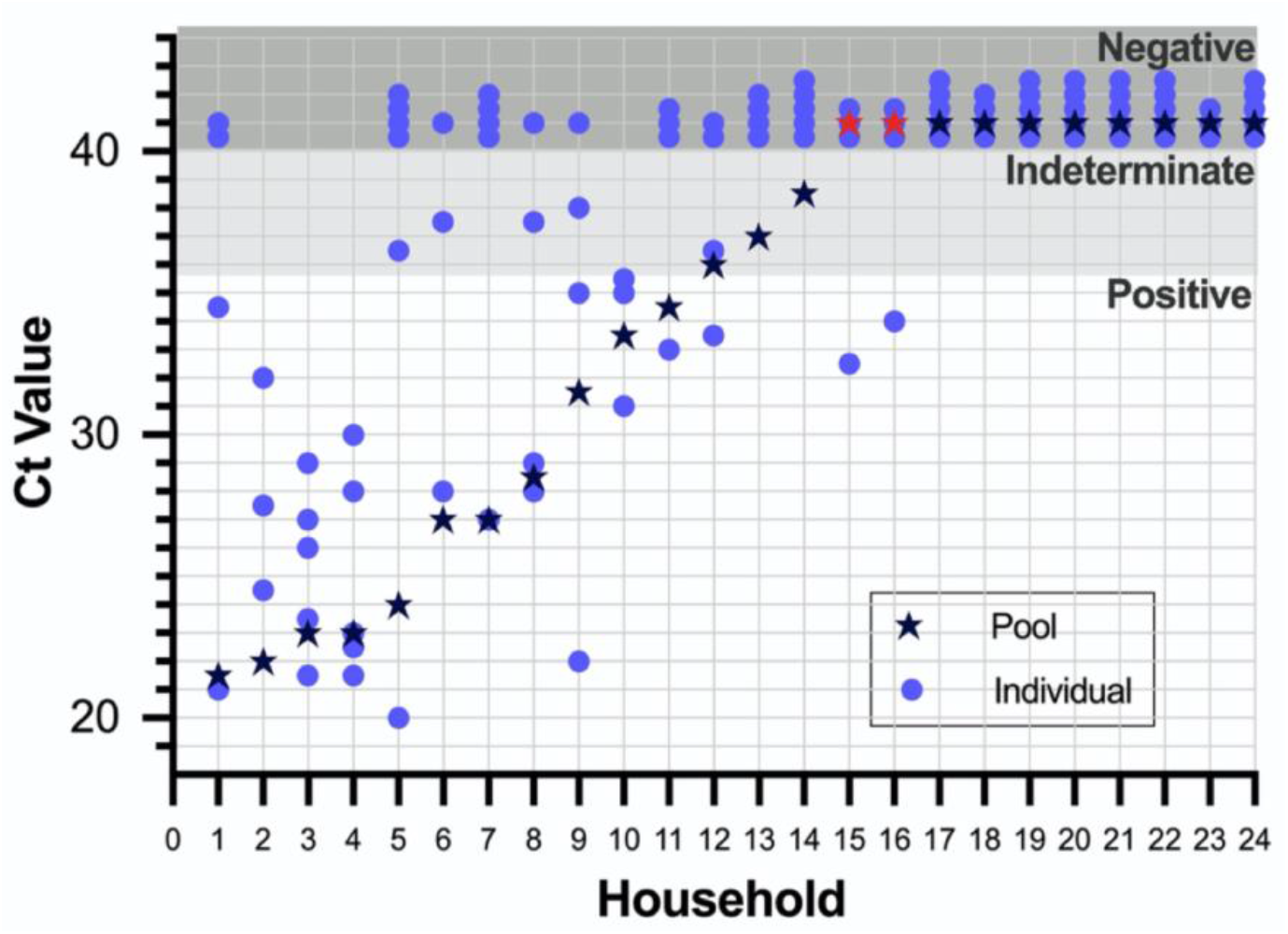
Comparison of Ct values from nasal mid-turbinate (NMT) swabs pooled from households of 3-5 persons at the point of care vs. concurrently collected individual NMT swabs. Among the pools collected from 24 households (listed along the x axis in order of decreasing viral loads), 2 pools with discordant results from individual swabs are depicted as red stars. Viral loads derived from the Ct values for each sample and the corresponding pool are found in Table S2.

Similarly, when individual sample lysates were pooled in the lab at varying pool sizes, none of the 2 sets of experimental pools containing a sample with a viral load of 10^1 copies/uL were positive (**Figure 5**). Of the 3 sets of pools containing a sample with a viral load of 10^2 copies/uL, 2 were positive at every pool size, while the remaining set was positive within pools of 5 and 10 samples, but indeterminate when the pool size was increased to 15 and 20 samples. The remaining pools constructed with samples with a viral load >10^2 copies/uL were positive across all pool sizes. The average total Ct value increase for the pools that remained positive at a pool size of 20 samples was 5.1 cycles, close to the expected 4.3 cycle increase for a sample diluted 1:20 using a PCR with 100% amplification efficiency.

**Figure 5.**
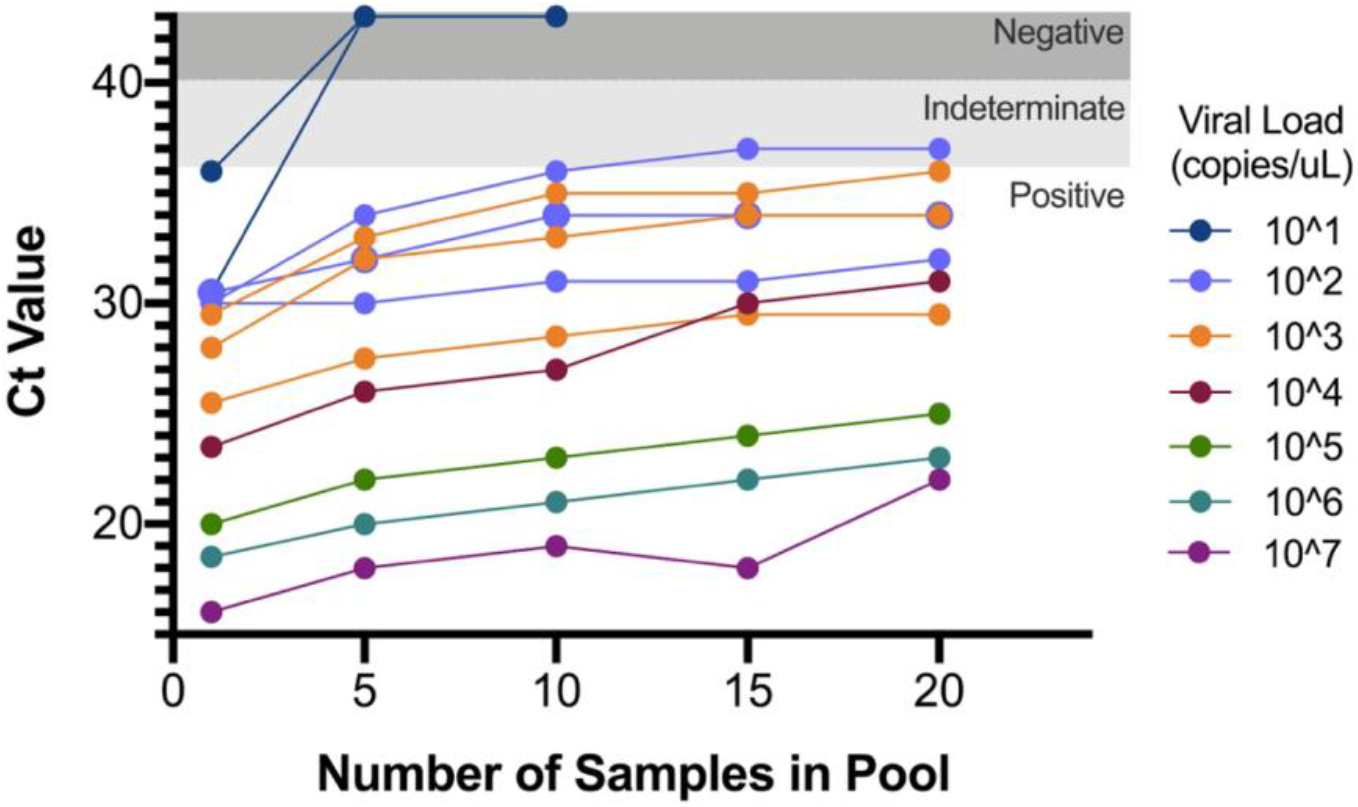
Ct values of increasing pool sizes constructed containing a single positive sample with varying viral loads. Viral transport media from a single positive sample with a viral load ranging from 10^1 to 10^7 were used to construct pool sizes of 5, 10, 15, 20.

## DISCUSSION

In a highly exposed outpatient cohort, we found nasal swabs to be reasonably sensitive, capturing 87% of SARS-CoV-2 infections diagnosed by nasopharyngeal sampling. This estimate is similar to most other outpatient studies showing >85% concordance between self-collected nasal swabs (either nasal mid-turbinate or anterior nasal swabs) and clinician-collected nasopharyngeal sampling (2,10–13). Not all studies are consistent however, likely due to heterogeneity in testing environments, and inclusion of non-acute samples collected during follow-up (14,15).

By calculating quantitative SARS-CoV-2 viral loads, our study gives clarity on where sensitivity is diminished (16). For the majority of participants in which nasal sampling failed to detect virus, the NP viral load was <1000 copies/uL, at a level that is likely non-infectious. Of these participants, 7/11 were antibody-positive at the time of sampling (unpublished data), and for the 8/11 participants still reporting symptoms, the average duration of reported symptoms was 6.5 days. Thus nasal samples are likely adequate for clinical diagnosis of acute infections to help expand testing capacity, but insensitivity to low viral load infections should be taken into consideration. On average, the decreased sensitivity of NMT swabs led to a little over a log decrease in viral copies/uL compared to NP swabs.

Our pragmatic approach of “show one, then do one” meant that nasal swabs were both clinician and self-collected. Also, since we often collected two nasal swabs per person, one from each nostril, our sampling strategy may have slightly underperformed relative to other studies that sample both nostrils with the same swab. It should be noted that we tested flocked and 3D-printed lattice swabs, but did not test dry swabs or non-flocked cotton swabs. Where high quality swabs are not available, but other swab types are plentiful, a strategy of combining oral and nasal samples appears promising (17).

3D-printed plastic swabs may also help address supply chain shortages (18,19). We first acquired prototype NMT lattice swabs from Resolution Medical in anticipation of shortage of supplies for our research study. In our limited testing, the prototype 3D-printed NMT lattice swabs showed high categorical concordance with NP swabs and also yielded similar viral loads compared to flocked NMT swabs. Similar high concordance has been demonstrated for 3D-printed nasopharyngeal swabs (18–20). Anecdotally, the prototype 3D-printed were observed to be more uncomfortable for study participants compared to flocked NMT swabs, a sentiment shared by other studies (18). This may have contributed to the higher proportion of samples deemed as inadequate sampling.

Labs also face VTM shortages requiring alternate transport media (21,22). Reagents which can inactivate virus and also keep samples stable at ambient temperature may be particularly apt substitutes (23). We used 1xDNA/RNA shield (Zymogen), an RNA preservation agent that has been widely used to inactivate SARS-CoV-2 and other respiratory viruses in various sample types and is now part of saliva and NMT Shield collection kits that have received FDA emergency use authorization (24–26). In our hands, storage of nasal swab samples in Shield did not improve their overall diagnostic sensitivity, but positive NMT swabs stored in Shield maintained quantitative viral loads more similar to those detected in concurrently collected NP swabs.

Pooling specimens in the lab is a well-documented strategy to accelerate SARS-CoV-2 testing in high-throughput settings (4–6). As in previous studies, we found that although Ct values do increase with pooling, the strategy can be broadly successful (27–31). Samples with viral loads at or near the limit of detection (31), or <10^3^ viral copies/uL in the CDC EUA assay we adopted, may go undetected as pool sizes increase. This was even more apparent when pooling swabs at the point of collection, which we piloted as unsupervised self-collection of 3D-printed swabs into the same conical tube containing 5mL of VTM.

Our findings add to the evidence base for nasal swabs as an adequate substitute for PCR-based clinical diagnosis of SARS-CoV-2 infection in outpatient settings where nasopharyngeal sampling is challenging. Viral recovery can be maintained even when immediate cold chain is not possible by storing swabs in an RNA preservation agent that also deactivates infectious virus. Combined with pooling specimens in the lab, these practical strategies can help expand testing in resource-constrained settings.

## Data Availability

Data are available upon request

## ACKNOWLEDGMENTS

We thank the whole Co-HOST study team, the IDEEL lab, as well as our wonderful study participants. This work was supported by funds from the UNC Department of Medicine, the UNC COVID-19 Response Fund/Health Foundation, and the National Center for Advancing Translational Sciences (NCATS), National Institutes of Health (UL1TR002489). DW was supported by the North Carolina Policy Collaboratory at UNC with funding from the North Carolina Coronavirus Relief Fund established and appropriated by the North Carolina General Assembly. Prototype 3D-printed NMT swabs were provided by Resolution Medical.

