## Supplementary Material for "Practical strategies for SARS-CoV-2 RT-PCR testing in resource-constrained settings"

**Table S1:** Results of RT-qPCRs across nCoVPC plasmid control concentrations

| Target descriptive | Level concentrations (viral copies/uL) | Number of Positive Hit Counts | Total Test Counts |
|---|---|---|---|
| N1 | 100,000 | 26 | 26 |
| N1 | 50,000 | 3 | 3 |
| N1 | 20,000 | 33 | 33 |
| N1 | 2,000 | 33 | 33 |
| N1 | 200 | 33 | 33 |
| N1 | 20 | 33 | 33 |
| N1 | 10 | 20 | 21 |
| N1 | 2 | 11 | 33 |
| N2 | 100,000 | 26 | 26 |
| N2 | 50,000 | 3 | 3 |
| N2 | 20,000 | 33 | 33 |
| N2 | 2,000 | 33 | 33 |
| N2 | 200 | 33 | 33 |
| N2 | 20 | 33 | 33 |
| N2 | 10 | 18 | 21 |
| N2 | 2 | 17 | 33 |

**Table S2: qRT-PCR results for pooled 3D-NMT swabs and concurrently collected individual flocked NMT swabs.** Pools, and their comprising individuals, are denoted by shaded rows. Individuals of the household who were not available for pooling are marked as not applicable. 24 pools and 106 individual swabs were collected.

| Family ID | Subject ID | Ct value | Avg viral RNA copies/uL (NMT) | Pool result +/- | Ct Value of Pool | Average viral RNA copies/uL of pool |
|---|---|---|---|---|---|---|
| COV004 D21 | COV004 | undet | 0 | - | undet | 0 |
| | COV004-001 | undet | 0 | | | |
| | COV004-002 | undet | 0 | | | |
| | COV004-004 | undet | 0 | | | |
| COV012 D1 | COV012 | 29 | 2,439 | + | 23 | 54,917 |
| | COV012-001 | 27 | 7,196 | | | |
| | COV012-002 | 21.5 | 144,769 | | | |
| | COV012-003 | 26 | 8,223 | | | |
| | COV012-004 | 23.5 | 55,478 | | | |
| COV017 D1 | COV017 | 28 | 3,603 | + | 23 | 99,275 |
| | COV017-001 | 23 | 135,846 | | | |
| | COV017-002 | 30 | 518 | | | |
| | COV017-003 | 22.5 | 162,938 | | | |
| | COV017-004 | 21.5 | 481,149 | | | |
| COV020 D22 | COV020 | undet | 0 | indet | 38.5 | 4 |
| | COV020-001 | undet | 0 | | | |
| | COV020-002 | undet | 0 | | | |
| | COV020-003 | undet | 0 | | | |
| | COV020-004 | 39.5 | 3 | | | |
| COV021 D14 | COV021 | undet | 0 | - | undet | 0 |
| | COV021-001 | undet | 0 | | | |
| | COV021-002 | undet | 0 | | | |
| | COV021-003 | undet | 0 | | | |
| | COV021-004 | undet | 0 | | | |
| COV021 D21 | COV021 | undet | 0 | - | undet | 0 |
| | COV021-001 | undet | 0 | | | |
| | COV021-002 | undet | 0 | | | |
| | COV021-003 | undet | 0 | | | |
| | COV021-004 | undet | 0 | | | |
| COV026 D7 | COV026 | undet | 0 | - | undet | 0 |
| | COV026-001 | undet | 0 | | | |
| | COV026-002 | undet | 0 | | | |
| | COV026-003 | undet | 0 | | | |
| | COV026-004 | undet | 0 | | | |
| COV026 D14 | COV026 | undet | 0 | - | undet | 0 |
| | COV026-001 | undet | 0 | | | |
| | COV026-002 | undet | 0 | | | |
| | COV026-003 | undet | 0 | | | |
| | COV026-004 | undet | 0 | | | |
| COV027 D7 | COV027 | 31 | 298 | + | 33.5 | 34 |
| | COV027-001 | 35 | 24 | | | |
| | COV027-002 | 35 | 21 | | | |
| | COV027-003 | 35.5 | 16 | | | |
| COV027 D14 | COV027 | undet | 0 | indet | 36 | 7 |
| | COV027-001 | 36.5 | 7 | | | |
| | COV027-002 | 33.5 | 31 | | | |
| | COV027-003 | undet | 0 | | | |
| COV029 D8 | COV029 | undet | 0 | + | 31.5 | 131 |
| | COV029-001 | 38 | 5 | | | |
| | COV029-002 | 35 | 29 | | | |
| | COV029-003 | 22 | 134,705 | | | |
| COV030 D8 | COV030 | 29 | 1532 | + | 28.5 | 2183 |
| | COV030-001 | undet | 0 | | | |
| | COV030-002 | 28 | 2767 | | | |
| | COV030-003 | 37.5 | 5 | | | |
| COV030 D22 | COV030 | undet | 0 | indet | 37 | 3 |
| | COV030-001 | undet | 0 | | | |
| | COV030-002 | undet | 0 | | | |
| | COV030-003 | undet | 0 | | | |
| COV032 D7 | COV032 | 32.5 | 66 | - | undet | 0 |
| | COV032-001 | undet | 0 | | | |
| | COV032-002 | undet | 0 | | | |
| | COV032-003 | undet | 0 | | | |
| COV032 D21 | COV032 | 34 | 24 | - | undet | 0 |
| | COV032-001 | undet | 0 | | | |
| | COV032-002 | undet | 0 | | | |
| | COV032-003 | undet | 0 | | | |
| COV033 D7 | COV033 | undet | 0 | - | undet | 0 |
| | COV033-001 | undet | 0 | | | |
| | COV033-002 | undet | 0 | | | |
| | COV033-003 | undet | 0 | | | |
| | COV033-004 | undet | 0 | | | |
| COV041 D7 | COV041 | undet | 0 | + | 24 | 28,967 |
| | COV041-001 | 20 | 371,312 | | | |
| | COV041-002 | 36.5 | 4 | | | |
| | COV041-003 | undet | 0 | | | |
| | COV041-004 | undet | 0 | | | |
| | COV041-005 | undet | 0 | | | |
| COV045 D7 | COV045 | undet | 0 | - | undet | 0 |
| | COV045-001 | undet | 0 | | | |
| | COV045-002 | undet | 0 | | | |
| | COV045-003 | n/a | n/a | | | |
| COV049 D7 | COV049 | 32 | 607 | + | 22 | 9,468 |
| | COV049-001 | 27.5 | 59,704 | | | |
| | COV049-002 | 27.5 | 19,719 | | | |
| | COV049-003 | 24.5 | 892,519 | | | |
| COV051 D9 | COV051 | undet | 0 | - | undet | 0 |
| | COV051-001 | undet | 0 | | | |
| | COV051-002 | undet | 0 | | | |
| | COV051-003 | undet | 0 | | | |
| | COV051-004 | undet | 0 | | | |
| COV052 D1 | COV052 | 21 | 270,034 | + | 21.5 | 152,869 |
| | COV052-001 | undet | 0 | | | |
| | COV052-002 | undet | 0 | | | |
| | COV052-003 | 34.5 | 22 | | | |
| COV052 D8 | COV052 | 33 | 69 | + | 34.5 | 14 |
| | COV052-001 | undet | 0 | | | |
| | COV052-002 | undet | 0 | | | |
| | COV052-003 | undet | 0 | | | |
| COV055 D8 | COV055 | undet | 0 | + | 27 | 6,152 |
| | COV055-001 | 37.5 | 9 | | | |
| | COV055-002 | 28 | 3,800 | | | |
| | COV055-005 | n/a | n/a | | | |
| COV059 D8 | COV059 | undet | 0 | + | 27 | 6,417 |
| | COV059-001 | 27 | 8,367 | | | |
| | COV059-002 | undet | 0 | | | |
| | COV059-003 | undet | 0 | | | |
| | COV059-004 | undet | 0 | | | |

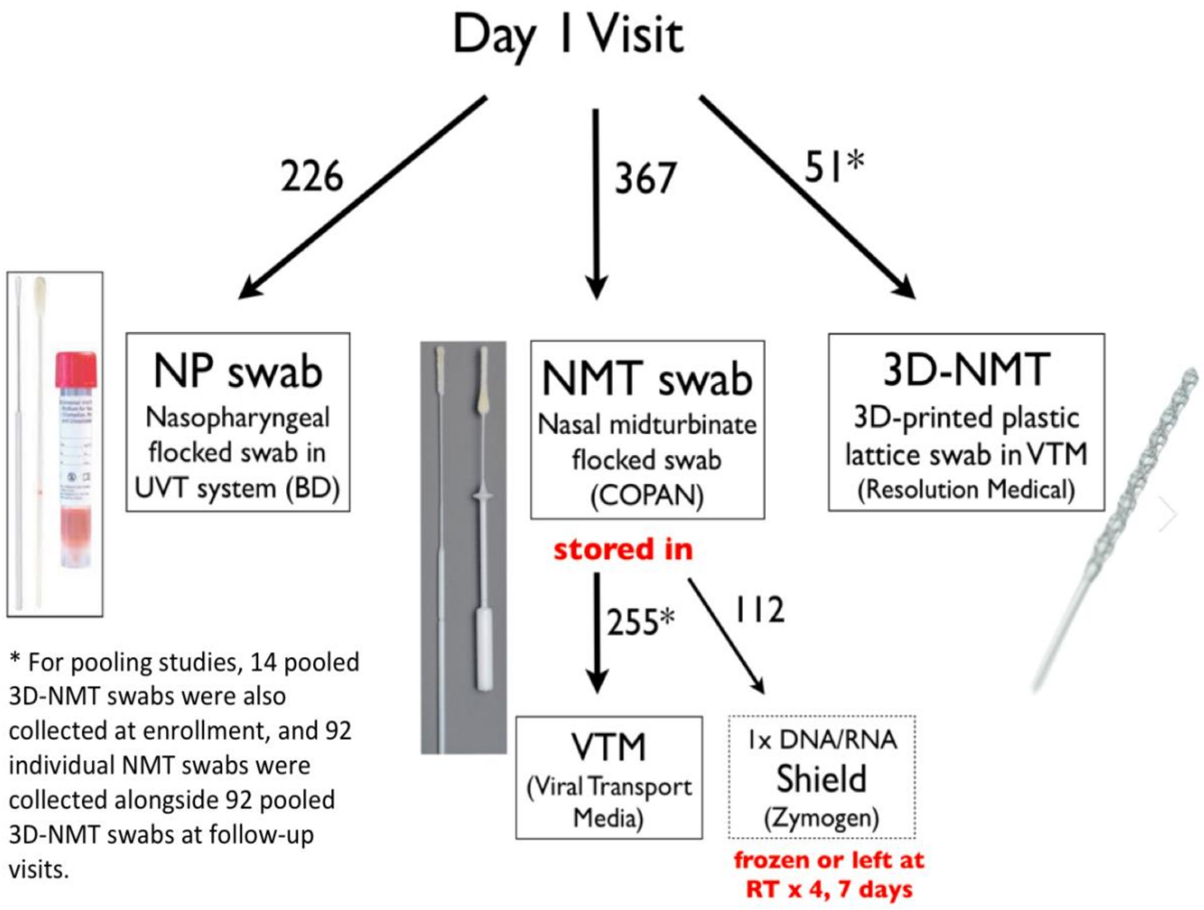


**Figure S1. Schematic of swab collection strategies at enrollment.** For each study participant at enrollment, a nasopharyngeal (NP) swab was collected as the gold standard diagnostic test. Up to 2 nasal midturbinate swabs - flocked COPAN swabs (NMT) and/or 3D-printed plastic lattice swabs (3D-NMT) - were concurrently collected, one from each nostril, for comparison. 3D-printed swabs were autoclaved before use per manufacturer's instructions

Swabs were either stored in viral transport media (VTM) or 1x DNA/RNA Shield (1xShield). Samples stored in 1xShield were further analyzed via aliquots that were either immediately frozen (D1 Shield) or left out at room temperature for 4 or 7 days (D4 Shield, D7 Shield) before storage at -80°C. A total of 620 individual swabs were collected at enrollment, while 184 swabs were collected to test pooling at follow-up visits.

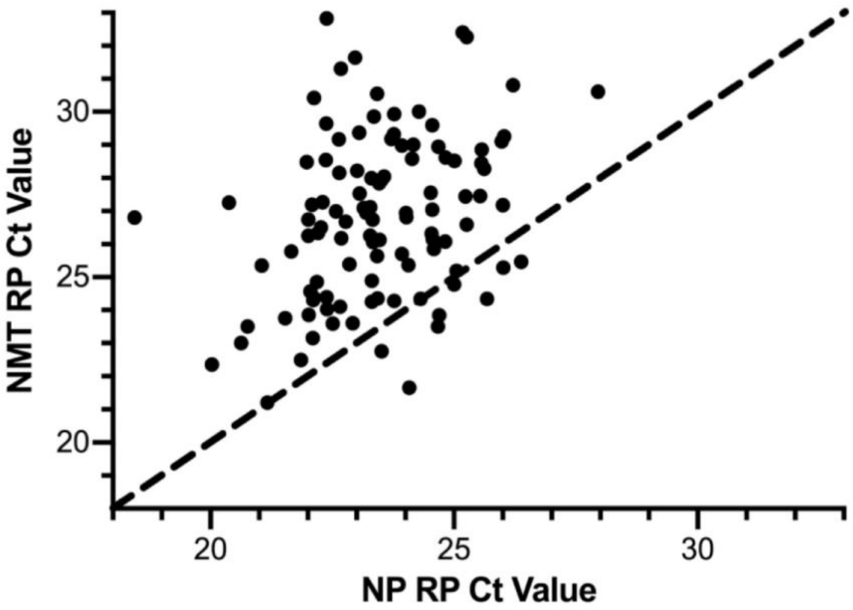


**Figure S2. Comparison of human RPP30 gene (RP) cycle threshold (Ct) values from paired nasopharyngeal (NP) and nasal mid-turbinate (NMT) swabs.** Higher RP Ct values were observed in NMT swabs compared to NP swabs with an average Ct value difference of 3.1 cycles, suggesting better sampling. A y=x line is drawn for reference.
